# Smoking and Vaping Among a National Sample of U.S. Adults During the COVID-19 Pandemic

**DOI:** 10.1101/2021.03.18.21253902

**Authors:** Sara Kalkhoran, Douglas E. Levy, Nancy A. Rigotti

**Affiliations:** Division of General Internal Medicine, Department of Medicine, Massachusetts General Hospital, Boston, MA, USA; Tobacco Research and Treatment Center, The Mongan Institute, Massachusetts General Hospital; Harvard Medical School, Boston, MA, USA

## Abstract

**Introduction:** With concerns about cigarette smoking being a risk factor for severe disease from COVID-19, understanding nicotine and tobacco use patterns is important for preventive efforts. We aimed to understand changes in product use behaviors among U.S. adult combustible cigarette smokers and electronic cigarette (e-cigarette) users.

**Methods:** In August 2020, we conducted a cross-sectional survey of a nationally-representative sample of adults age >18 in NORC’s AmeriSpeak Panel who reported past 6-month use of combustible cigarettes or e-cigarettes. Multivariable logistic regression assessed factors associated with increased product use and quit attempts since hearing about COVID-19.

**Results:** 1024 past 6-month cigarette smokers and/or e-cigarette users were surveyed. Among cigarette smokers, 45% reported no change in cigarette smoking and 33% increased cigarette smoking since hearing about COVID-19. Higher stress was associated with increased cigarette smoking. Among e-cigarette users, 41% reported no change in and 23% reported increasing e-cigarette use. 26% of cigarette smokers and 41% of e-cigarette users tried to quit because of COVID-19. Higher perceived risk of COVID-19 was associated with attempts to quit combustible cigarettes (AOR 2.37, 95% CI 1.59-3.55) and e-cigarettes (AOR 3.14, 1.73-5.70).

**Conclusions:** Cigarette and e-cigarette use patterns varied in response to the COVID-19 pandemic. Most cigarette smokers and e-cigarette users perceived product use as increasing COVID-19-related health risks, and this was associated with attempts to quit. Others, especially those reporting higher stress, increased product use. Proactive provision of cessation support to smokers and e-cigarette users may help mitigate stress-related increases in product use during the COVID-19 pandemic.

## Introduction

As cases of coronavirus disease 2019 (COVID-19) continue to increase in the United States and worldwide, the management of potentially modifiable factors associated with the risk of severe disease or death is an important component of preventive efforts. Health organizations have identified smoking as a risk factor for severe disease from SARS-COV-2 infection,^1,2^ an association that has also been suggested by several,^3,4,5–7^ but not all,^8^ systematic reviews. Data on the association between electronic cigarette (e-cigarette) use (vaping) and COVID-19 outcomes is lacking, although there has been some concern about vaping being associated with increased COVID-19 risk among young people.^9^

Many smokers turn to cigarettes when they are stressed. Stressful life events have been negatively associated with quitting smoking,^10^ and perceived stress has been associated with nicotine withdrawal symptoms.^11^ Therefore, some smokers and e-cigarette users may increase their product use during the uncertain time of a pandemic. At the same time, other smokers might decrease their tobacco use because they have chronic heart or lung disease that they perceive puts them at a higher risk of negative outcomes from COVID-19, or because they have trouble obtaining or using their normal products during business closures or shelter-in-place orders. Studies are urgently needed to understand changes in product use and behaviors among people who smoke or vape during the COVID-19 pandemic. A survey of dual users of cigarettes and e-cigarettes in the U.S. done early in the pandemic found that between 20-25% of participants had tried to quit cigarettes or e-cigarettes to reduce harm from COVID-19.^12^ On the other hand, approximately 30% reported increasing their use of cigarettes and e-cigarettes. More recent data from larger, nationally-representative groups of smokers and e-cigarette users are needed to further understand these use patterns.

Better understanding of smokers’ and e-cigarette users’ behaviors and attitudes is important for designing targeted outreach and education for this population. We conducted a cross-sectional survey of combustible cigarette smokers and/or e-cigarette users from a nationally-representative sample of U.S. adults to assess: (1) changes in product use as a result of COVID-19, (2) factors associated with increased product use, and (3) factors associated with product quit attempts.

## Methods

### Design and Participants

We conducted a cross-sectional survey of a nationally-representative sample of adults in the National Opinion Research Center’s (NORC) AmeriSpeak Panel.^13^ For the Amerispeak Panel, U.S. households are sampled using probability-and address-based sampling methods to cover approximately 97% of the U.S. household population. Sampled households are contacted for potential participation in the Amerispeak Panel through mail, telephone, and field interviews, and informed consent is provided by participants at the time of registration. Most panel members participate in surveys online, and households that are not connected to the internet can take surveys by telephone. Panel members participate in surveys approximately 2-3 times a month. For the present study, eligibility criteria included age 18 or older, English-speaking, and use of either combustible cigarettes or electronic nicotine products/e-cigarettes since February 2020 (i.e. past 6-month use). A screening questionnaire was used to identify past 6-month cigarette and e-cigarette users; e-cigarette users and dual users were sampled separately from cigarette-only users to obtain adequate sample sizes for questions related to e-cigarette use. Only de-identified data was obtained from NORC, and this study was deemed exempt by the Institutional Review Board at Mass General Brigham. Surveys were conducted from August 20 to August 25, 2020.

### Measures

Participants reporting past 6-month use of combustible cigarettes or e-cigarettes were asked whether they currently used the products every day, some days, or not at all and how many years they had been using the products. Dual use was defined as use of both combustible cigarettes and e-cigarettes in the past 6 months.

With respect to COVID-19 and change in product use, past 6-month combustible cigarette smokers were asked “Since hearing about the coronavirus (COVID-19), would you say the amount you are smoking has…” with answer choices (1) increased a lot, (2) increased a little, (3) stayed about the same, (4) decreased a little, or (5) decreased a lot. For regression models, this was dichotomized into “increased” and “not increased”. Current combustible cigarette smokers who were still smoking at the time of the survey reported whether hearing about COVID-19 affected their interest in reducing or stopping smoking (Yes, less interested; Yes, more interested; or No, not affected), and whether hearing about COVID-19 has led them to try to stop smoking (Yes/No). We defined trying to quit smoking as reporting (1) that COVID led them to try to stop smoking or (2) that they had completely stopped smoking since hearing about COVID-19. Past 6-month e-cigarette users were asked the same questions about use of e-cigarettes/vaping, and dual users of cigarettes and e-cigarettes were asked these questions about both products. Past 6-month combustible cigarette smokers were asked their opinion about smoking and their risk of getting COVID-19 or having a more serious case in any way (“smoking definitely increases my risk”, “smoking might increase my risk”, “smoking does not affect my risk”, “smoking might decrease my risk”, “smoking definitely decreases my risk”). Past 6-month e-cigarette users/vapers were asked the same question about vaping and COVID risk.

All participants were asked whether they had been tested for COVID-19 or had been told by a medical professional that they were infected with COVID-19. Potential stressors were assessed by asking participants about whether COVID-19 changed how much time they spend working and how much they worry about having financial problems as a result of the COVID-19 pandemic (on a 5-point scale from “not at all” to “very much”). Participants also completed the perceived stress scale 4 (PSS-4),^14^ which is scored from 0-16, with higher levels indicating higher stress. This scale is not specific to COVID-19 but was asked of participants during the COVID-19 pandemic.

Sociodemographic factors included age, gender, race/ethnicity, level of education, and region of residence.

### Statistical analysis

First, we described cigarette and e-cigarette use behaviors and perceived risks related to COVID-19. Then, we conducted univariable logistic regression analyses assessing predictors of (1) increased cigarette smoking or vaping due to COVID-19, (2) trying to quit cigarette smoking or vaping, or (3) increased interest in reducing or stopping cigarette smoking or vaping among participants who were still using the products at the time of the survey. Variables with p<0.15 in univariable analyses were included in multivariable logistic regression models with sociodemographic characteristics and cigarette or e-cigarette use. Only participants with complete data on study variables were included in the multivariable models. All analyses adjusted for complex survey procedures and sample weights using Stata version 13 (StataCorp LLC, College Station, Texas). All n’s are reported as unweighted and all percentages are weighted.

## Results

Overall, 4,734 of the 20,455 (23.1%) invited Amerispeak Panel members completed the screening questionnaire. Of these individuals, 678 (14.3%) reported either e-cigarette use or dual use of e-cigarettes and cigarettes in the past 6 months and 549 (11.6%) reported cigarette smoking only during this time and were therefore eligible for the survey. Among those eligible, 507 (74.7%) of the e-cigarette or dual users and 517 (94.3%) of the cigarette smokers completed the survey. Among those in the e-cigarette sample, 336 reported vaping e-cigarettes only, and 171 reported both smoking cigarettes and vaping e-cigarettes. Characteristics of the study population by past 6-month use of e-cigarettes and cigarettes in shown in Table 1.

**Table 1.** Characteristics of smokers reporting past 6-month cigarette smoking or e-cigarette use/vaping.

|  | Past 6-month smokers<br>(n=688)<sup>a</sup> | Past 6-month e-cigarette<br>users/vapers (n=507)<sup>a</sup> |
| --- | --- | --- |
| Age in years, mean (SD) | 45 (1.0) | 36 (0.8) |
| Female gender | 54% (350) | 43% (233) |
| Race/ethnicity |  |  |
| NH White | 66% (466) | 57% (288) |
| NH Black | 11% (100) | 15% (88) |
| Hispanic | 15% (69) | 22% (95) |
| Other | 8% (53) | 7% (36) |
| Level of education |  |  |
| HS or less | 53% (215) | 51% (153) |
| Some college | 27% (340) | 29% (228) |
| 4-year degree or more | 19% (133) | 20% (126) |
| Region |  |  |
| Northeast | 18% (116) | 13% (68) |
| Midwest | 25% (212) | 20% (136) |
| South | 38% (220) | 42% (156) |
| West | 18% (140) | 26% (147) |
| Cigarette smoking |  |  |
| Past 6-month cigarette smoking | 100% (688) | 34% (171) |
| Current cigarette smoking | 81% (553) | 27% (128) |
| Current daily cigarette smoking | 56% (393) | 18% (80) |
| E-cigarette use |  |  |
| Past 6-month e-cigarette use | 29% (171) | 100% (507) |
| Current e-cigarette use | 21% (124) | 67% (336) |
| Current daily e-cigarette use | 6% (39) | 17% (95) |
| Duration of cigarette smoking (current users) | 22 years (0.9) | 16 years (1.4) |
| Duration of vaping (current users) | 2.5 years (0.3) | 2 years (0.1) |
| Total stress score (0-16), mean (SD) | 6.5 (0.2) | 6.9 (0.2) |
| COVID affected finances | 67% (436) | 77% (378) |
| Have had to use savings | 29% (187) | 30% (149) |
| Had to borrow money/get loan | 21% (121) | 20% (94) |
| Could not make bill payments | 20% (116) | 23% (107) |
| Cut down on spending for food | 27% (182) | 32% (154) |
| Needed to cut down on expenses in general | 45% (303) | 40% (208) |
| Mean number of finances affected | 1.4 (0.1) | 1.4 (0.1) |
| Worry about financial problems<br>(1 – not at all, 5 – very much) | 2.8 (0.1) | 2.9 (0.1) |
| COVID affected work hours |  |  |
| Lost job | 13% (79) | 14% (82) |
| Yes, working less | 14% (83) | 22% (91) |
| Yes, working more | 7% (35) | 11% (47) |
| No, working same | 33% (240) | 34% (168) |
| No, not working before COVID | 33% (248) | 19% (112) |
| Tested for COVID | 27% (135) | 38% (171) |
| Diagnosed with COVID | 2% (11) | 8% (34) |
| <sup>a</sup>Groups are not mutually exclusive |  |  |

### Changes in Product Use Since COVID-19

Among cigarette smokers who had smoked since hearing about COVID-19 (n=601), 45% reported no change in cigarette smoking, 21% reported decreased cigarette smoking, and 33% reported increased cigarette smoking (Figure 1a). These proportions did not differ significantly between cigarette-only smokers and dual users of e-cigarettes and cigarettes (p=0.30, data not shown in tables). Among past 6-month cigarette smokers who reported quitting prior to the COVID-19 pandemic (n=85), 15% reported relapsing back to cigarette smoking. In multivariable analyses, factors associated with increased cigarette smoking among participants who had smoked since hearing about COVID-19 were higher perceived stress, greater worry about financial problems due to the COVID-19 pandemic, and working more during the pandemic (Table 2).

**Figure 1.**
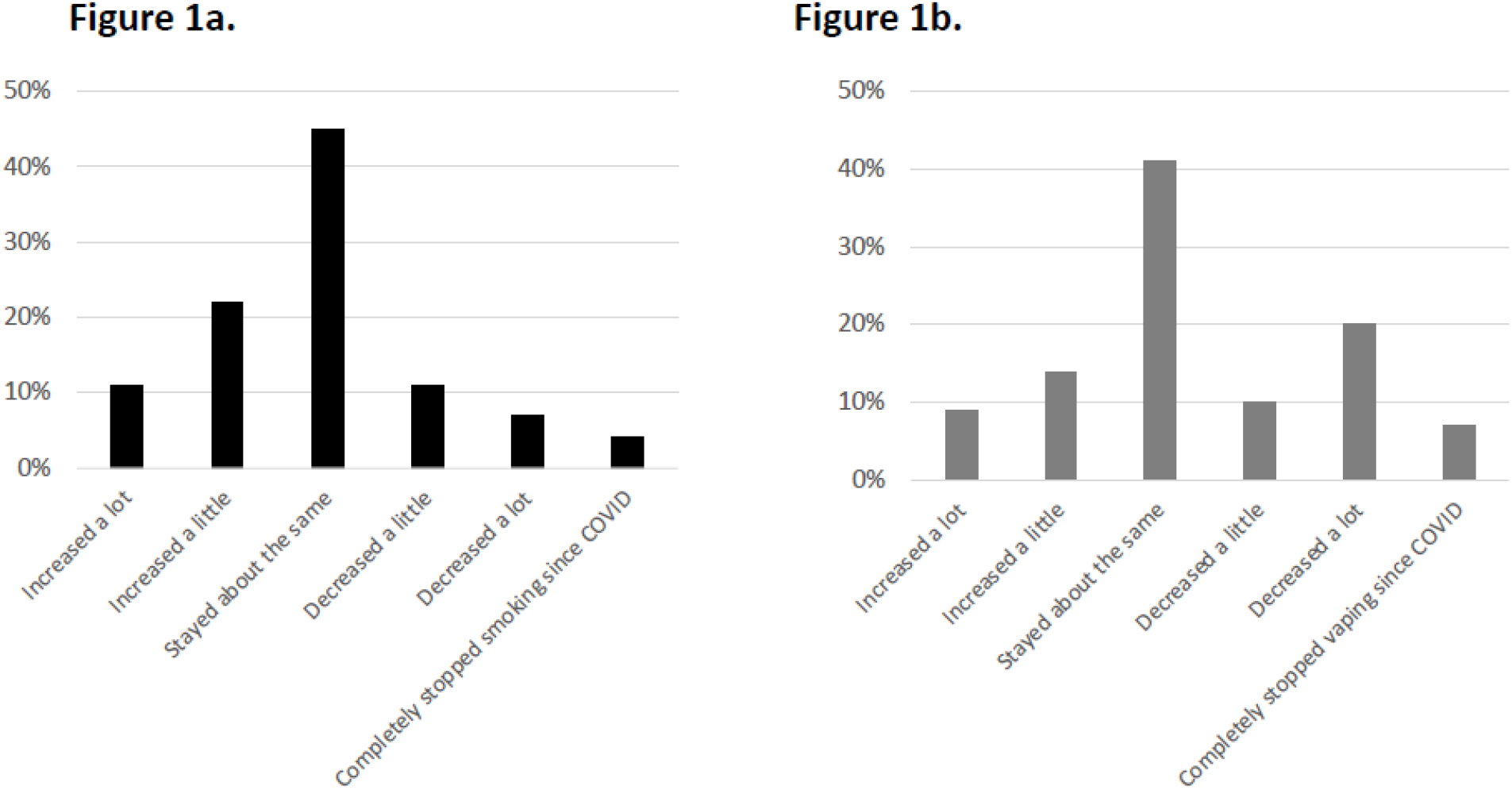
(a) Cigarette smoking behaviors among past 6-month cigarette smokers who reported having smoked cigarettes since hearing about COVID-19. (b) E-cigarette use/vaping behavior among past 6-month e-cigarette users who reported having vaped since hearing about COVID-19.

**Table 2.** Predictors of increased smoking or vaping due to COVID-19.

|  | Increased smoking due to COVID-19<br>(n=596) |  | Increased vaping due to COVID-19<br>(n=378) |  |
| --- | --- | --- | --- | --- |
|  | OR | aOR | OR | aOR |
| Age | 0.97 (0.96-0.99) | 0.99 (0.97-1.01) | 1.01 (0.99-1.03) | 1.01 (0.99-1.03) |
| Female gender | 1.15 (0.68-1.95) | 1.09 (0.66-1.81) | 1.79 (0.99-3.23) | **1.96 (1.08-3.54)** |
| White race | 0.70 (0.40-1.23) | 0.89 (0.52-1.54) | 0.84 (0.47-1.51) | 0.97 (0.54-1.75) |
| Education |  |  |  |  |
| HS or less | 1.69 (0.91-3.15) | 1.13 (0.59-2.17) | 0.58 (0.28-1.18) | 0.60 (0.27-1.33) |
| Some college | 1.18 (0.67-2.09) | 0.78 (0.41-1.47) | 0.88 (0.47-1.66) | 0.69 (0.31-1.50) |
| 4-year degree or more | ref | ref | ref | ref |
| Region of residence |  |  |  |  |
| Northeast | Ref | Ref | Ref | Ref |
| Midwest | 1.13 (0.61-2.12) | 1.15 (0.56-2.35) | **0.33 (0.13-0.87)** | **0.36 (0.13-0.96)** |
| South | 0.54 (0.28-1.04) | 0.48 (0.22-1.05) | **0.23 (0.10-0.56)** | **0.27 (0.11-0.68)** |
| West | 1.19 (0.50-2.86) | 0.89 (0.39-2.02) | 0.61 (0.25-1.48) | 0.56 (0.23-1.38) |
| Past 6-month cigarette smoking | -- | -- | 1.13 (0.62-2.09) | 1.23 (0.65-2.33) |
| Past 6-month e-cigarette use | 1.23 (0.71-2.12) | 1.19 (0.67-2.13) | -- | -- |
| Total stress score (0-16) | **1.23 (1.13-1.33)** | **1.15 (1.06-1.26)** | **1.11 (1.01-1.21)** | 1.09 (0.99-1.20) |
| Worry about financial problems<br>(1 – not at all, 5 – very much) | **1.62 (1.32-1.98)** | **1.31 (1.04-1.67)** | 1.14 (0.90-1.43) | -- |
| COVID affected work hours |  |  |  |  |
| Working less | **2.05 (1.20-3.49)** | 1.28 (0.69-2.36) | **1.95 (1.05-3.62)** | 1.82 (0.96-3.42) |
| Working more | **5.58 (1.60-19.43)** | **2.98 (1.06-8.34)** | 0.61 (0.23-1.62) | 0.70 (0.26-1.89) |
| No change | Ref | Ref | Ref | Ref |
| Perceived risk of COVID from<br>smoking/vaping (1 – definitely<br>decreases, 5 – definitely<br>increases) | 1.19 (0.93-1.53) | -- | **1.42 (1.01-1.99)** | 1.28 (0.91-1.78) |

**Figure 2.**
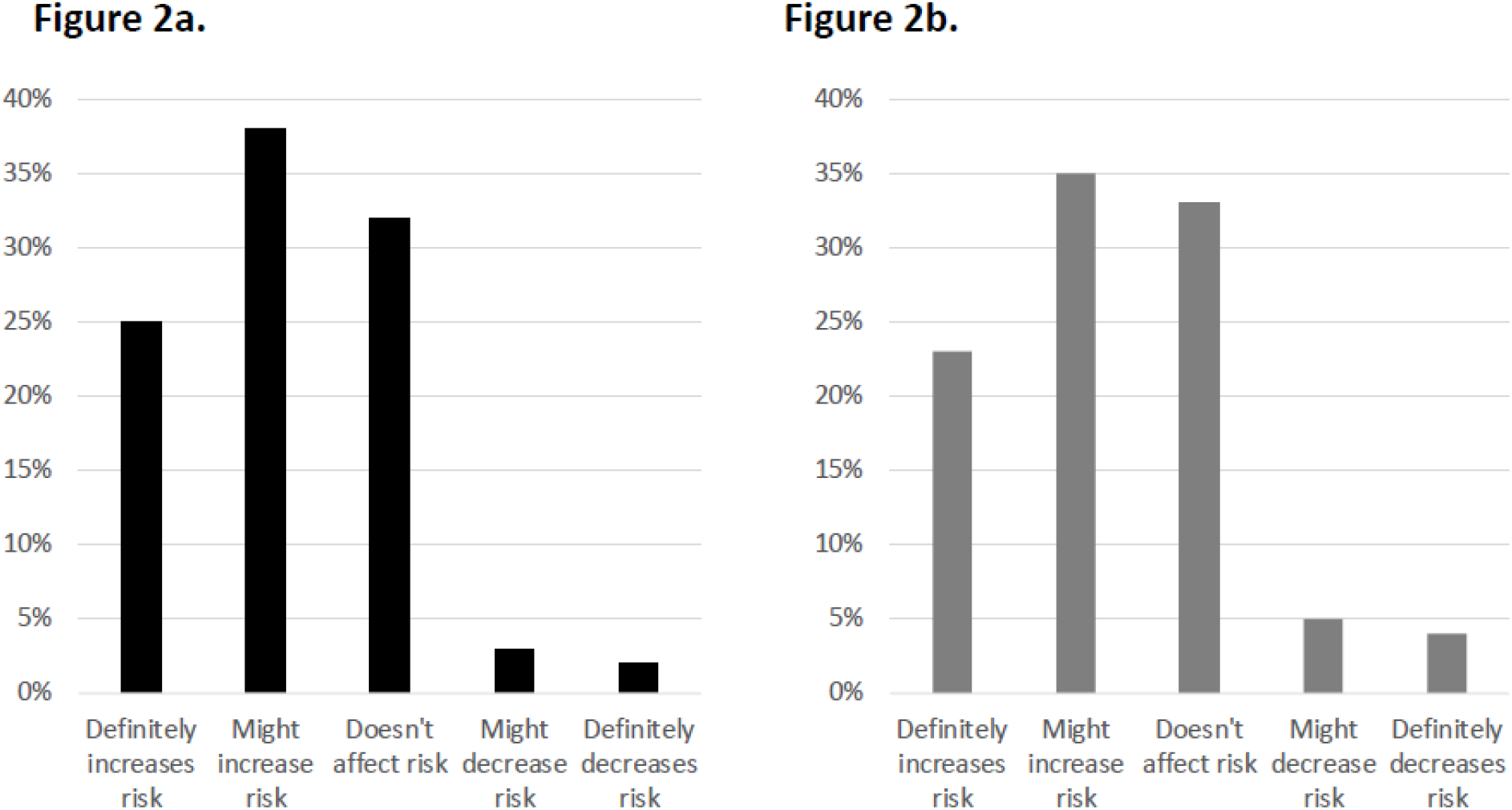
(a) Opinion about cigarette smoking and risk of getting COVID-19 or having a more serious outcome among past 6-month cigarette smokers. (b) Opinion about e-cigarette use/vaping and risk of getting COVID-19 or having a more serious outcome among past 6-month e-cigarette users/vapers.

Among e-cigarette users who had vaped since hearing about COVID-19 (n=384), 41% reported no change in e-cigarette use, 37% reported decreased e-cigarette use, and 23% reported increased e-cigarette use (Figure 1b). E-cigarette-only users and dual users did not differ significantly on this measure (p=0.85). Among the past 6-month e-cigarette users who reported quitting prior to the COVID-19 pandemic (n=113), most (82%) reported staying quit while 18% reported relapsing back to e-cigarette use. In multivariable analyses, increased vaping since hearing about COVID-19 was associated with female gender and negatively associated with residing in the Midwest or South (vs Northeast), but associations with COVID-19 factors (financial stress, work hours, COVID-19 risk) were not significant (Table 2).

### Perceived Risk of COVID-19 Due to Smoking and Vaping

Most (63%) past 6-month cigarette smokers perceived that cigarette use “might increase” or “definitely increases” their risk of contracting COVID-19 or having a more serious outcome, while 32% did not think cigarette smoking affected their risk (supplemental Figure). Perceived risk of COVID-19 did not differ significantly between cigarette-only smokers and dual users (p=0.22, data not shown in tables). Similarly, most (59%) past 6-month e-cigarette users reported feeling that e-cigarette use “might increase” or “definitely increases” their risk of COVID-19 or a more serious outcome, while one-third (33%) did not think vaping affected their risk (supplemental Figure). E-cigarette-only users and dual users did not differ significantly in perceived risk of COVID-19 (p=0.89).

### Quit Attempts and Interest in Quitting Smoking or Vaping

Among participants who had smoked cigarettes since hearing about COVID-19, 26% reported trying to quit because of COVID-19. Dual users did not differ significantly from cigarette-only users in trying to quit (p=0.07). The belief that smoking increases risk of COVID-19 or having a more severe case was associated with trying to quit cigarette smoking, and daily smoking and more years of cigarette smoking were negatively associated with trying to quit (Table 3). Among participants who had smoked cigarettes since hearing about COVID-19 and were still smoking at the time of the survey, 35% reported increased interest in quitting smoking and this did not differ significantly between dual users and cigarette-only users (p=0.90). The belief that smoking increases risk of COVID-19 or having a more severe case was associated with increased interest in quitting or reducing cigarette smoking (AOR 2.82, 95% CI 1.98-4.02), while daily smoking was negatively associated with increased interest in quitting or reducing cigarette smoking (AOR 0.35, 0.19-0.63, Supplemental Table 1).

**Table 3.** Predictors of trying to quit smoking or vaping due to COVID-19.

|  | Tried to quit smoking because of COVID-19 (n=557) |  | Tried to quit vaping because of COVID-19 (n=301) |  |
| --- | --- | --- | --- | --- |
|  | OR | OR | OR | aOR |
| Age | 1.00 (0.98-1.02) | **1.04 (1.02-1.07)** | 0.99 (0.97-1.02) | 0.99 (0.96-1.02) |
| Female gender | 1.05 (0.62-1.80) | 1.48 (0.83-2.62) | 1.24 (0.68-2.25) | 1.35 (0.63-2.89) |
| White race | 0.61 (0.34-1.08) | 0.63 (0.35-1.14) | 0.63 (0.34-1.15) | 0.75 (0.33-1.73) |
| Education |  |  |  |  |
| HS or less | 1.14 (0.59-2.20) | 2.34 (0.95-5.75) | 1.25 (0.60-2.58) | 1.74 (0.69-4.37) |
| Some college | 0.97 (0.53-1.78) | 1.69 (0.71-4.03) | 0.72 (0.36-1.44) | 0.75 (0.31-1.80) |
| 4-year degree or more | ref | ref | ref | ref |
| Region of residence |  |  |  |  |
| Northeast | Ref | Ref | Ref | Ref |
| Midwest | 1.66 (0.83-3.29) | 2.19 (1.03-4.66) | 0.75 (0.30-1.86) | 0.62 (0.20-1.93) |
| South | 1.89 (0.96-3.69) | 1.94 (0.87-4.35) | 0.82 (0.34-2.01) | 0.67 (0.22-2.06) |
| West | 1.44 (0.52-3.94) | 1.41 (0.51-3.87) | 1.01 (0.40-2.54) | 0.71 (0.20-2.49) |
| Past 6-month cigarette smoking | -- | -- | 0.59 (0.31-1.13) | **0.45 (0.21-0.96)** |
| Past 6-month e-cigarette use | 1.73 (0.95-3.15) | 1.68 (0.76-3.69) | -- | -- |
| Duration of cigarette smoking | **0.98 (0.96-0.99)** | **0.97 (0.95-0.99)** | -- | -- |
| Duration of e-cigarette use |  |  | 0.93 (0.75-1.14) | -- |
| Daily cigarette smoking | **0.21 (0.12-0.37)** | **0.21 (0.11-0.38)** | -- | -- |
| Daily e-cigarette use | -- | -- | **0.21 (0.09-0.45)** | **0.23 (0.10-0.55)** |
| Total stress score (0-16) | 1.02 (0.96-1.09) | -- | 1.09 (0.99-1.20) | 1.05 (0.92-1.19) |
| Worry about financial problems (1 – not at all, 5 – very much) | 1.12 (0.93-1.34) | -- | 1.19 (0.95-1.51) | 0.99 (0.71-1.37) |
| COVID affected work hours |  |  |  |  |
| Working less | 1.21 (0.65-2.26) | -- | 1.67 (0.87-3.17) | 1.32 (0.55-3.15) |
| Working more | 1.18 (0.32-4.33) |  | 1.34 (0.50-3.60) | 0.99 (0.32-3.09) |
| No change | Ref |  | Ref | ref |
| Perceived risk of COVID from smoking/vaping (1 – definitely decreases, 5 – definitely increases) | **2.17 (1.45-3.23)** | **2.37 (1.59-3.55)** | **2.31 (1.48-3.60)** | **3.14 (1.73-5.70)** |

Among participants who had vaped since hearing about COVID-19, 41% reported trying to quit vaping because of COVID-19. Dual users did not differ significantly from e-cigarette-only users in trying to quit (p=0.11). The belief that vaping increases risk of COVID-19 or having a more severe case was associated with trying to quit vaping, while daily e-cigarette use and past 6-month cigarette smoking were negatively associated with trying to quit vaping (Table 3). Among participants who used e-cigarettes since hearing about COVID-19 and were still vaping at the time of the survey, 43% reported increased interest in quitting vaping and this did not differ significantly between dual users and e-cigarette-only users (p=0.53). The belief that vaping increases risk of COVID-19 or having a more severe case was associated with increased interest in quitting or reducing cigarette vaping (AOR 2.14, 95% CI 1.48-3.09) as was working more during the pandemic (AOR 4.01, 1.32-12.21), while daily e-cigarette use was negatively associated with increased interest in quitting or reducing cigarette smoking (AOR 0.25, 0.12-0.54, Supplemental Table 1).

## Discussion

Among cigarette smokers and e-cigarette vapers in a nationally-representative panel of U.S. adults, we found that product use patterns and behaviors varied in response to the COVID-19 pandemic. Approximately a third of combustible cigarette smokers and a quarter of e-cigarette users increased their product use during the pandemic, and personal and economic stressors were associated with increased use among combustible cigarette smokers. At the same time, 26% and 41% of combustible cigarette smokers and e-cigarette users, respectively, reported trying to quit product use during the pandemic. Most cigarette smokers and e-cigarette users perceived their product use as increasing their risk of COVID-19 or a more serious outcome. Higher perceived risk related to COVID-19 was associated with attempts to quit both cigarettes and e-cigarettes, while daily product use was negatively associated with quit attempts. The study findings highlight the range of behavioral responses that users of addictive products have had in response to an ongoing pandemic.

We found that 33% of combustible cigarette smokers and 23% of e-cigarette users increased their product use, similar to prior studies.^12,15^ Prior studies have found that while some e-cigarette users had concerns about product access during the COVID-19 pandemic, many were still able to obtain the products either through stockpiling of products, online purchasing, or from their usual channels either through vape shop non-compliance with or circumventing of business closure orders.^16–18^ We found factors associated with various forms of stress – total stress, worry about financial problems, and increased working hours - to be associated with increased combustible cigarette smoking during the COVID-19 pandemic. A study of current smokers in the UK during the COVID-19 pandemic also found an association between psychosocial and mental health factors and changes in smoking consumption.^19^ These data are consistent with findings from non-pandemic times in which high stress levels have been associated with continued cigarette smoking, and the association between changes in perceived stress and changes in smoking status.^20^ Targeted outreach to smokers during the pandemic to assist with stress management and avoidance of smoking escalation can help to mitigate some of these stress-related increases in smoking consumption. This is particularly important as tobacco use continues to be the leading preventable cause of death worldwide.

Increased interest in quitting product use was reported by 35% of combustible cigarette smokers and 43% of e-cigarette users during the COVID-19 pandemic, consistent with other, non-representative samples of U.S. adults surveyed earlier in the pandemic.^12,15^ Therefore, the pandemic may be a time of increased receptiveness to help for smoking or vaping cessation. A study in the UK found higher odds of trying to quit and of quitting among past-year smokers after a COVID-19 lockdown, and while smokers were no more likely to use evidence-based smoking cessation treatment after the lockdown, use of remote smoking cessation support in the form of telephone support, websites, or telephone applications increased during this time.^22^ Bolstering access to tobacco cessation resources that do not require in-person contact may be one way for smokers and vapers to act on their interest in quitting and successfully quit.

In line with our findings related to perceived risk of COVID-19 and attempts to quit, prior work by Grummon et al. using a convenience sample of U.S. adults found that both messaging related to traditional harms of smoking and those specifically related to COVID-19 had higher perceived effectiveness for discouraging smoking compared to control messages.^21^ In contrast, while messaging about traditional harms of vaping were associated with higher perceived effectiveness for discouraging vaping compared to control, the same was not seen for COVID-19 related messages.^21^ Thus, education of smokers on the relationship between smoking and COVID-19 risk may serve as an important motivator for cessation and/or reducing potential increases in product use. Further research is necessary to assess the effects of public health messaging on smoking and vaping during the pandemic and optimize messaging content for these individuals.

This study is subject to several limitations. First, data are cross-sectional and therefore longitudinal trends in combustible cigarette and e-cigarette use could not be assessed. Second, all data are self-reported, including retrospective reports of use patterns, and therefore subject to recall bias. Third, questions about risk perceptions combined perceived risk of getting COVID-19 and having a more serious case, so the answers to the individual components of this question cannot be ascertained. Finally, data were collected at the end of August 2020, when COVID-19 cases were lower compared to Spring 2020 and Fall 2020, and therefore participant behaviors may not be reflective of behaviors at other times during the pandemic.

In conclusion, this survey of a large nationally-representative sample of cigarette smokers and e-cigarette users in the U.S. revealed a range of product use behaviors in response to the COVID-19 pandemic, with some trying to quit product use and others increasing their product use. Most combustible cigarette smokers and e-cigarette users perceived higher COVID-19-related health risks as a result of their product use, factors that appeared to be associated with attempts to quit product use. Outreach to smokers and e-cigarette users to provide cessation assistance during this time may help to support quit attempts and reduce stress-related increases in product use.

## Data Availability

Data reported in the manuscript are available from the authors.

## Acknowledgments

This study was supported by the National Heart, Lung, and Blood Institute (NHLBI) of the National Institutes of Health (K23HL136854 to Dr. Kalkhoran). The funder had no role in the design and conduct of the study; collection, management, analysis, and interpretation of the data; preparation, review, or approval of the manuscript; and decision to submit the manuscript for publication.

## References

1. World Health Organization. Smoking and COVID-19. Accessed November 24, 2020. https://www.who.int/publications-detail-redirect/WHO-2019-nCoV-Sci_Brief-Smoking-2020.2

2. CDC. Coronavirus Disease 2019 (COVID-19). Centers for Disease Control and Prevention. Published February 11, 2020. Accessed November 24, 2020. https://www.cdc.gov/coronavirus/2019-ncov/need-extra-precautions/people-with-medical-conditions.html

3. Simons D, Shahab L, Brown J, Perski O. The association of smoking status with SARS-CoV-2 infection, hospitalization and mortality from COVID-19: a living rapid evidence review with Bayesian meta-analyses (version 7). Addiction. Published online October 2, 2020. doi:10.1111/add.15276

4. Gülsen A, Yigitbas BA, Uslu B, Drömann D, Kilinc O. The Effect of Smoking on COVID-19 Symptom Severity: Systematic Review and Meta-Analysis. Pulm Med. 2020;2020:7590207. doi:10.1155/2020/7590207

5. Patanavanich R, Glantz SA. Smoking Is Associated With COVID-19 Progression: A Meta-analysis. Nicotine Tob Res. 2020;22(9):1653–1656. doi:10.1093/ntr/ntaa082

6. Karanasos A, Aznaouridis K, Latsios G, et al. Impact of Smoking Status on Disease Severity and Mortality of Hospitalized Patients With COVID-19 Infection: A Systematic Review and Meta-analysis. Nicotine Tob Res. 2020;22(9):1657–1659. doi:10.1093/ntr/ntaa107

7. Reddy RK, Charles WN, Sklavounos A, Dutt A, Seed PT, Khajuria A. The effect of smoking on COVID-19 severity: A systematic review and meta-analysis. J Med Virol. Published online August 4, 2020. doi:10.1002/jmv.26389

8. Lippi G, Henry BM. Active smoking is not associated with severity of coronavirus disease 2019 (COVID-19). Eur J Intern Med. 2020;75:107–108. doi:10.1016/j.ejim.2020.03.014

9. Gaiha SM, Cheng J, Halpern-Felsher B. Association Between Youth Smoking, Electronic Cigarette Use, and COVID-19. J Adolesc Health. 2020;67(4):519–523. doi:10.1016/j.jadohealth.2020.07.002

10. McKee SA, Maciejewski PK, Falba T, Mazure CM. Sex differences in the effects of stressful life events on changes in smoking status. Addiction. 2003;98(6):847–855. doi:10.1046/j.1360-0443.2003.00408.x

11. Lawless MH, Harrison KA, Grandits GA, Eberly LE, Allen SS. Perceived stress and smoking-related behaviors and symptomatology in male and female smokers. Addict Behav. 2015;51:80–83. doi:10.1016/j.addbeh.2015.07.011

12. Klemperer EM, West JC, Peasley-Miklus C, Villanti AC. Change in tobacco and electronic cigarette use and motivation to quit in response to COVID-19. Nicotine Tob Res. Published online April 28, 2020. doi:10.1093/ntr/ntaa072

13. About Amerispeak. Accessed May 8, 2020. http://amerispeak.norc.org/about-amerispeak/Pages/default.aspx

14. Cohen S, Kamarck T, Mermelstein R. A global measure of perceived stress. Journal of health and social behavior. Published online 1983:385–396.

15. Streck JM, Kalkhoran S, Bearnot B, et al. Perceived risk, attitudes, and behavior of cigarette smokers and nicotine vapers receiving buprenorphine treatment for opioid use disorder during the COVID-19 pandemic. Drug and Alcohol Dependence. 2021;218:108438. doi:10.1016/j.drugalcdep.2020.108438

16. Soule EK, Mayne S, Snipes W, Guy MC, Breland A, Fagan P. Impacts of COVID-19 on Electronic Cigarette Purchasing, Use and Related Behaviors. Int J Environ Res Public Health. 2020;17(18). doi:10.3390/ijerph17186762

17. Berg CJ, Callanan R, Johnson TO, et al. Vape shop and consumer activity during COVID-19 non-essential business closures in the USA. Tobacco Control. Published online October 18, 2020. doi:10.1136/tobaccocontrol-2020-056171

18. Gaiha SM, Lempert LK, Halpern-Felsher B. Underage Youth and Young Adult e-Cigarette Use and Access Before and During the Coronavirus Disease 2019 Pandemic. JAMA Netw Open. 2020;3(12):e2027572. doi:10.1001/jamanetworkopen.2020.27572

19. Tzu-Hsuan Chen D. The psychosocial impact of the COVID-19 pandemic on changes in smoking behavior: Evidence from a nationwide survey in the UK. Tob Prev Cessat. 2020;6. doi:10.18332/tpc/126976

20. Cohen S, Lichtenstein E. Perceived stress, quitting smoking, and smoking relapse.:14.

21. Grummon AH, Hall MG, Mitchell CG, et al. Reactions to messages about smoking, vaping and COVID-19: two national experiments. Tob Control. Published online November 2020. doi:10.1136/tobaccocontrol-2020-055956

22. Jackson SE, Garnett C, Shahab L, Oldham M, Brown J. Association of the COVID-19 lockdown with smoking, drinking and attempts to quit in England: an analysis of 2019–20 data. Addiction. n/a(n/a). doi:https://doi.org/10.1111/add.15295

